# Multi-Omic Profiling Reveals Epigenetic Drivers of Immunotherapy Resistance in Multiple Myeloma

**DOI:** 10.1101/2025.04.15.25325788

**Authors:** Larissa Haertle, Natalia Buenache Cuenda, Francisco J. Villena González, Isabel Espejo Díaz, Paula Lazaro del Campo, Juan Manuel Rosa-Rosa, Rafael Alberto Alonso Fernandez, Maria Nieves Lopez Muñoz, José María Sánchez-Pina, Almudena García-Ortiz, Antonio Valeri, Santiago Barrio, Fátima Al-Shahrour, Florian Bassermann, Joaquín Martinez-Lopez, Tomás Di Domenico

## Abstract

Targeted immunotherapies against B-cell maturation antigen (BCMA) have transformed the treatment landscape of Multiple Myeloma (MM). Fc receptor-like 5 (FCRL5) has emerged as an alternative target. However, resistance frequently emerges within months, posing a significant clinical challenge. Structural alterations and mutations in *BCMA* only account for the minority of cases and insights into BCMA antigen escape remain largely unknown. This study investigates novel (epi)genetic mechanisms of antigen escape through comprehensive multi-omic Oxford Nanopore profiling of sequential pre-treatment and relapse samples. We identify acquired DNA-hypermethylation across the entire *BCMA* gene, and hypermethylation of the *FCRL5* promoter, both resulting in epigenetic gene silencing as novel resistance mechanisms through which MM cells evade therapy. These findings underscore the dynamic clonal evolution of MM under therapeutic pressure and highlight the critical role of epigenetic modifications in resistance. Furthermore, we demonstrate the potential of advanced sequencing technologies for capturing epigenetic and complex genomic alterations in clinical settings, paving the way for personalized treatment strategies and predictive biomarkers for early resistance detection.

**Statement of significance:** Acquired DNA hypermethylation of *BCMA* and *FCRL5* regulatory regions, leading to gene expression downregulation, represent novel epigenetic resistance mechanisms to anti-BCMA and anti-FCRL5 immunotherapies. Furthermore, DNA methylation marks serve as a molecular memory of therapeutic pressure, capturing the treatment history of cancer cells.

## Introduction

Several of the most potent immunotherapies currently used to treat Multiple Myeloma (MM) target the B-cell maturation antigen (BCMA) through T-cell mediated elimination of the tumor. These include BCMA chimeric antigen receptor T-cells (CAR T) such as idecabtagene vicleucel (ide-cel), ciltacabtagene autoleucel (cilta-cel), or anitocabtagene autoleucel (anito-cel)^1–3^. Additional modalities include antibody-drug conjugates (ADC) like belantamab mafodotin (BLMF)^4^, as well as BCMAxCD3 bispecific T-cell engagers (TCEs) such as teclistamab, elranatamab, linvoseltamab, AMG420, AMG701, or TNB-383B^5^. Given the favourable safety profiles and efficacy of these therapies demonstrated in phase 1/2/3 clinical trials for relapsed or refractory MM patients (RRMM), ongoing trials investigate the inclusion of anti-BCMA therapies in earliest lines of treatment^3,6^. However, recent real-world data and updates of clinical trials have provided longer-term clinical follow-ups and exposed therapeutic limitations. One major clinical challenge remains innate and acquired resistance^3,6,7^. The percentage of non-responding and primary refractory patients varies widely depending on the specific therapy, trial, prior treatments, and patient characteristics, ranging from 2% to 69% with a significant number of responders experiencing disease progression within the first year and beyond^1,2,6^. Understanding the molecular mechanisms behind the transient durability of clinical response is challenging. MM is a disease with high genetic heterogeneity and complex clonal architecture. Frequent relapses and short remissions during the disease course are well-documented and associated with the expansion of genetic events^8–11^.

T cell-mediated selective pressure can drive antigen drift, altering the tumor’s antigenic landscape through the emergence of subclones with reduced or absent target antigens^6,12,13^. BCMA antigen loss, in particular, appears to be a key mechanism of MM cells evasion to anti-BCMA immunotherapies, as demonstrated with ide-cel^14,15^, teclistamab, elranatamab^7^, and linvoseltamab^9^. Large structural events on chromosome 16 and focal deletions of the *BCMA* locus were described as genetic events that, homozygously acquired, eliminate *BCMA* expression^3,7,9,14,15^. But also non-truncating missense mutations and in-frame deletions within the extracellular BCMA domain can impair the efficacy of anti-BCMA TCEs, despite detectable surface BCMA protein expression^3,7^. Consequently, the immunoselection of BCMA-negative or mutant clones represents a significant tumor-intrinsic driver of relapse. However, the described resistance alterations account for the minority of the clinically observed resistant cases, underscoring the presence of other unexplored resistance mechanisms. For instance, impaired FADD/BID signaling has been identified *in vitro* as a mediator of cross-resistance to BCMA-directed CAR T cells and teclistamab^16^. Tumor-extrinsic factors like immune exhaustion and a suppressive microenvironment also contribute to resistance^3^.

Given these challenges, alternative therapeutic targets such as GPRC5D and FCRL5 have garnered increasing interest. FCRL5, a homologous Fc receptor, predominantly expressed on B and plasma cells, has emerged as a promising target. Cevostamab, a bispecific FCRL5/CD3 antibody, has demonstrated efficacy in overcoming *BCMA* resistance. However, the mechanisms underlying resistance to FCRL5-targeting therapies remain largely unknown. Epigenetic alterations of the target gene may play a crucial role in acquired resistance. For instance, long-range epigenetic silencing of the *GPRC5D* promoter and enhancers, identified through ATAC-seq and bulk RNA-seq, was recently reported as an acquired resistance mechanism to talquetamab in two out of three patients studied^13^. Additionally, *GPRC5D* hypermethylation in transcriptional regulatory elements was observed in five MM patients who relapsed after GPRC5D CAR T-cell therapy^17^. Notably, DNA methylation alterations in *BCMA* and *FCRL5* have not yet been investigated. Given that DNA methylation is a key regulatory mechanism of gene expression through promoter/enhancer modifications and chromatin remodeling, understanding its role in resistance to these therapies is critical. DNA methylation is particularly important in B-cell development, tumorigenesis, and cancer progression, often occurring through localized hypo- or hypermethylation at regulatory CpG islands or transcription factor binding sites^18^. In previous work, we identified focal DNA hypermethylation of one *CRBN* enhancer as a resistance mechanism to immunomodulatory agents (IMiDs)^19^ and *PSMD5* promoter hypermethylation as a resistance mechanism to proteasome inhibitors (PIs)^8^, with respective prevalences of 67% and 25% in relapsed/refractory MM (RRMM) patients. This growing body of evidence underscores aberrant DNA methylation as a potentially key yet understudied driver of drug resistance.

Using Oxford Nanopore Technologies (ONT) long-read sequencing, here we conducted a comprehensive multi-omic analysis of sequential RRMM patient samples treated with anti-BCMA, anti-FCRL5, and anti-GPRC5D immunotherapies. Our approach simultaneously assessed single nucleotide variants (SNVs), copy number aberrations (CNAs), complex structural variants (SVs), whole-epigenome DNA methylation, and transcriptomic changes by RNA sequencing and flow cytometry. SNVs identified by ONT were technically validated using targeted tchDNA-Seq, while intrinsic *BCMA* methylation levels were assessed in immunotherapy-naive MM patients via Deep Bisulfite Sequencing (DBS), and in MM cell lines using pyrosequencing. This study reveals that acquired DNA hypermethylation silences *BCMA* and *FCRL5* expression, representing a novel epigenetic mechanism of resistance to targeted immunotherapies in MM.

## Methods

### Patient Selection

We collected PBMCs and sequential CD138+ purified primary tumor samples from two heavily pretreated MM patients pre- and post-immunotherapies (**Figure 1A**). No statistical method was used to predetermine the sample size. These patients were amongst the first to relapse after BCMA-targeted treatment at the H12O hospital Madrid, with sequential bone marrow aspirates available rich enough in tumor burden for parallel Nanopore sequencing, tchDNA-Seq and RNA-seq. Detailed treatment regimens and patient responses are included in **SI Figure 1**. Briefly, prior to sampling, all patients were quadruple-drug refractory and resistant to at least one IMiD (lenalidomide and/or pomalidomide), two PIs (bortezomib and carfilzomib), and an anti-CD38 antibody (daratumumab). Detailed patient characteristics are listed in **SI Table 1**. Resistance was defined as no response to the treatment or disease progression within 60 days of the last dose. Patient 1 was treated with the BCMA CAR T ARI0002h, cevostamab (FCRL5xCD3), talquetamab (GPRC5DxCD3) and elranatamab (BCMAxCD3) plus selinexor, a targeted small-molecule inhibitor of exportin-1 (XPO1). Patient 2 was treated with the BCMA ADC belantamab mafodotin (BLMF). The study was approved by the Clinical Research Ethics Committee (20/326) of the Biomedical Research Institute of the Hospital 12 de Octubre and conducted in accordance with the principles of the Declaration of Helsinki. All patients provided written informed consent.

**Figure 1.**
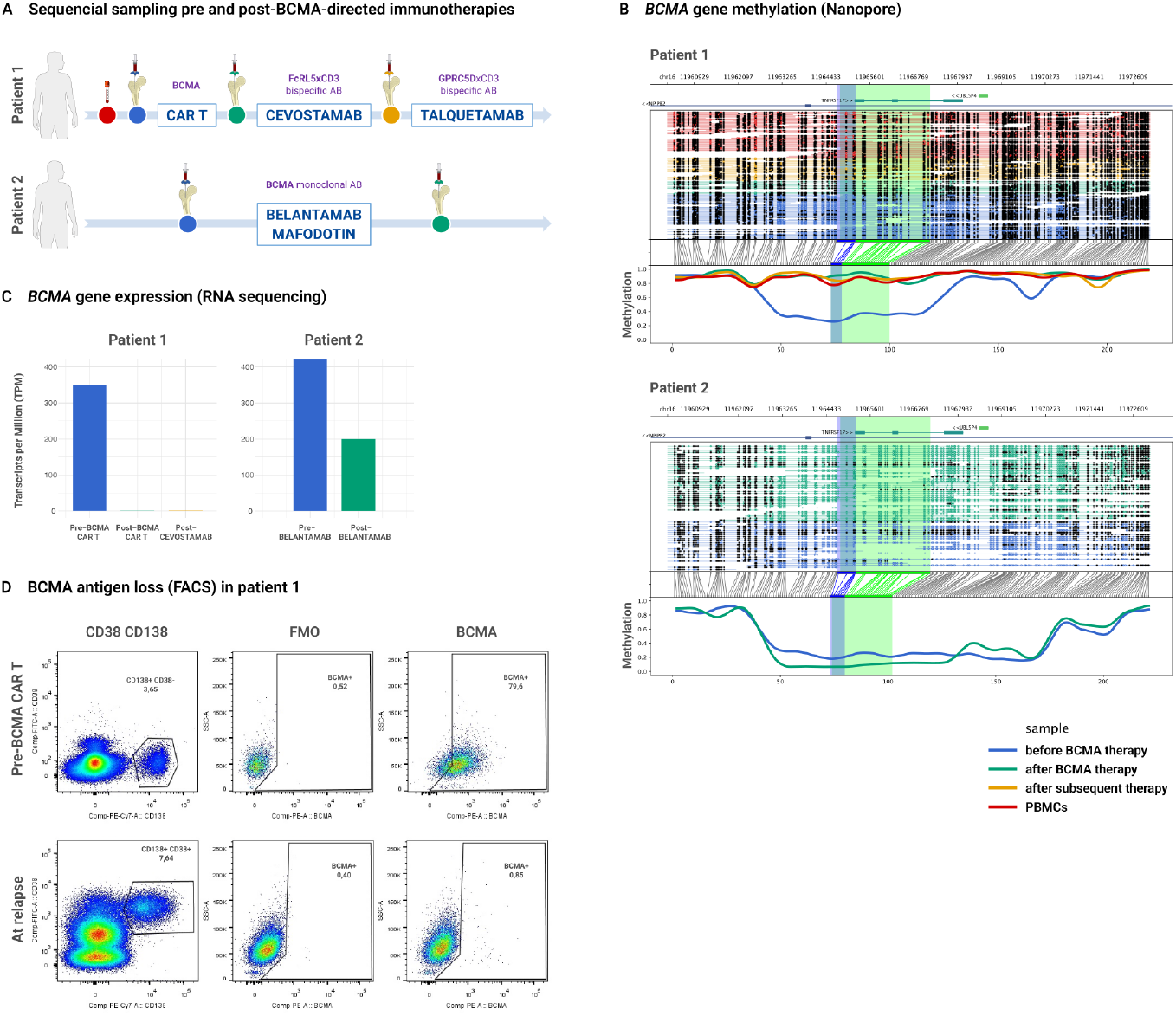
Acquired *BCMA* gene methylation after BCMA targeting immunotherapy and subsequent downregulation of the BCMA expression in patient 1. **(A)** Sequential sampling of CD138+ purified plasma cells from bone marrow aspirates of two RRMM patients before BCMA-targeting immunotherapy and at relapse. PBMCs served as controls. **(B)** Lollipop diagrams displaying *BCMA* methylation generated with the Methylartist suite. Coverage of the PBMC sample is indicated in red, pre-BCMA sample in blue, post-BCMA in green and subsequent sample in yellow. Regulatory regions defined by the Ensembl Regulatory Build are highlighted in coloured frames with promoter in blue and enhancer in green. The full *BCMA* gene is shown ±5 kB up- and downstream. The curve in the lower panel indicates the mean methylation per sample and CpG, ranging from 0 to 100% methylation. patient 1 acquired hypermethylation of the full *BCMA* gene in the relapse sample, which was absent in the pre-therapy sample. This hypermethylation was still detectable after subsequent cevostamab treatment. **(C)** Downregulation of BCMA gene transcripts after BCMA CAR T treatment in patient 1 and belantamab mafodotin in patient 2. (**D**) Subsequent antigen loss in patient 1. Dot plots of flow cytometry analysis of *BCMA* expression in MM plasma cells in patient 1, pre BCMA-CAR T cell therapy administration and at relapse.

### Sample Processing and Nanopore Sequencing

DNA and RNA were simultaneously extracted from MM cells using the AllPrep DNA/RNA/Protein Mini Kit (Qiagen). DNA concentration was measured with the Qubit dsDNA BR Assay kit (Thermo Fisher Scientific). Fragment length was assessed with the TapeStation 4200 (Agilent Technologies) instrument using the Genomic DNA Kit (Agilent Technologies). Average fragment length of all samples was 22 kB or longer. When possible, PBMCs were collected from the patients, and DNA was extracted using the Maxwell 48 RSC instrument (Promega) with the Maxwell RSC Whole Blood DNA Kit (Promega). Nanopore libraries were generated with, wherever possible, 1 µg of genomic DNA, however, half of the samples had lower concentrations, with the lowest being 268 ng. To achieve a whole genome-wide coverage of ≥ 20x, for samples with concentrations below 500 ng, after a 100-h runtime, the remaining sample was recovered from the flow cells (FCs) and rerun on a fresh FC following the ONT protocol “Library recovery from flow cells” (LIR_9178_v1_revl_11). Libraries were prepared with the Ligation Sequencing Kit V14 (ONT), the NEBNext Companion Module for ONT (New England Biolabs), and sequenced with R10.4.1 FCs (ONT) on the PromethION 2 Solo (ONT) device. The SQK-LSK114 guidelines were followed with some modifications: DNA shearing using the Covaris g-TUBEs was skipped, no extra DCS was used, LFB buffer was used instead of SFB to eliminate fragments smaller than 3 kb, pelleting and incubation times were prolonged to maximize recovery, and whatever was available but not more than 49 fmol were loaded onto a FC with an extra waiting time of 5 hours before starting the run. Detailed descriptions of the bioinformatic analysis of the Nanopore sequencing, the RNA sequencing, flow cytometry, DBS, pyrosequencing and tchDNA-Seq can be found in the **Supplemental Methods**. Primers are given in **SI Table 2**.

## Results

### Global hypomethylation in MM with acquired BCMA hypermethylation at relapse

Sampling was conducted as depicted in **Figure 1A:** for patient 1, at pre-BCMA CAR T, post-CAR T/pre-cevostamab, and post-cevostamab/pre-talquetamab; for patient 2, at pre- and post-BLMF. In general, global DNA hypomethylation was observed in plasma MM cells compared to PBMCs (**SI figure 2**). This aligns with prior literature indicating that MM is characterized by global hypomethylation, unlike other B-cell tumor malignancies^18^. However, when comparing solely pre- and post-treatment tumor samples, patient 1 exhibited increased methylation in the relapse sample. Notably, he acquired long-range *BCMA* hypermethylation at relapse following BCMA-directed immunotherapy (**Figure 1B**). The hypermethylated region (GRCh38 chr16:11962800-11969200) spanned 6,4 kb with 96 individual CpGs, encompassing the entire *BCMA* gene (chr16:1965210-11968068), which spans 2,86 kb and contains 40 CpGs and three epigenetically modified accessible regions (EMARs, chr16:11,964,344-11,964,555, chr16:11964697-11965522, chr16:11966138-11966679). For patient 1, *BCMA* methylation levels increased by 49%, rising from 41% pre-CAR T to 90% post-CAR T, and remained at 90% post-cevostamab. PBMCs showed methylation levels of 88%. Read-level analysis of methylation patterns revealed that this shift reflected clonal selection rather than gradual methylation acquisition (**SI Figure 7**). Pre-CAR T, sequencing reads segregated into two distinct patterns -one methylated and one unmethylated-indicating the coexistence of two clonal populations. Post-CAR T, all reads displayed a single, uniformly hypermethylated pattern, consistent with selective outgrowth of the methylated clone under therapeutic pressure. This homogeneous methylation pattern persisted post-cevostamab.

### Loss of BCMA expression correlates with BCMA hypermethylation

At relapse, patient 1 exhibited a loss of *BCMA* expression at both the transcriptional level (**Figure 1C**) and on the cell surface (**Figures 1C/D**). The transcript per million (TPM) value, initially 351.3 pre-BCMA CAR T, dropped to nearly undetectable levels post-treatment (0.9 post-CAR T and 1.4 post-cevostamab (**Figure 1C**). This transcriptional loss translated into antigen loss, as detected by FACS, where BCMA-positive cells decreased from 79.6% pre-CAR T to 0.85% at CAR T relapse (**Figure 1D**). Notably, in the sample collected before BCMA CAR T therapy, CD38 downregulation was observed on the cell surface, a phenomenon seen in patients treated with daratumumab^20,21^. Prior to sampling, the patient had received DKd as third-line therapy (**SI Figure 1A**). Importantly, *CD38* transcripts were detectable by RNA-seq at this timepoint (2.7 TPM), confirming that the absence of surface CD38 detection by flow cytometry does not reflect transcriptional silencing. *CD38* expression remained present across subsequent timepoints in both patients (**SI Figure 6**). FACS data was not available for the other patient. In patient 2, *BCMA* TPM values declined by half, from 423.7 pre-BLMF to 200.5 post-BLMF (**Figure 1C**). Importantly, none of the patients exhibited SNVs (**SI Table 3**) nor CNAs (**Figure 3, SI Figure 3, SI Table 4**) affecting the *BCMA* gene.

### BCMA methylation is absent in treatment-naive MM patients and MM cell lines

Using targeted Deep Bisulfite Sequencing (DBS), we analyzed the 5’promoter of *BCMA* (chr16:11965000-11965353, **SI Table 2**) in 14 MM patients naive to BCMA-directed therapy and eight PBMCs (**SI Figure 4**). All PBMCs showed methylation rates consistent with Nanopore sequencing results (mean: 70%±0.34, range: 49%-93%, **SI Figure 4**). In contrast, none of the MM patients showed notable *BCMA* methylation (mean: 2%±0.02, range: 0%-7%). These findings provide evidence that *BCMA* gene expression in mature B lymphocytes is regulated by DNA methylation. Furthermore, we screened five commonly used MM cell lines using two different pyrosequencing assays (**SI Table 2**). Since none of these cell lines harbored intrinsic *BCMA* methylation (mean methylation MM1S: 1.97%, NCI-H929: 1.03%, Amo1: 2.02%, OPM2: 2.13%, and KMS11: 2.21%, **SI Table 5**), assessing demethylating drugs such as 5-aza-2’-deoxycytidine and 5-azacytidine was not possible.

### FCRL5 promoter hypermethylation and epigenetic resistance memory of earlier treatments

In patient 1, *FCRL5* promoter methylation increased from 36% pre-CAR T to 54% post-CAR T and 52% post-cevostamab across a 6 kbp region (29 CpGs, chr1:157549000-157555000, **Figure 2B**). The hypermethylation was accompanied by a decline in *FCRL5* expression from 15.8 TPM to 3.9 TPM post-CAR T and 2.6 TPM post-cevostamab. The patient exhibited primary resistance to cevostamab (**SI Figure 1**) and was subsequently treated with talquetamab. Interestingly, screening for epigenetically modifying genes, we observed a moderate upregulation of the DNA methyltransferases DNMT1, DNMT3A and DNMT3B in the post-CAR T compared to the pre-CAR T sample in this patient (**Figure 2C**). He also harboured a hypermethylation in a 3’ regulatory region (chr12:121918000-121919401, 1.4 kbp, 65 CpGs, **Figure 2B**), overlapping with the *PSMD9* gene. Given the patient’s extensive prior exposure to multiple cycles of bortezomib and carfilzomib (**SI Figure 1**), a persistent methylation signature was observed at sampling time points. While PBMCs showed 49% methylation, all three sequential bone marrow samples exhibited >79% methylation, with complete loss of *PSMD9* gene expression (**Figure 2B**). Regrettably, there is no pre-PI sample of this patient available for comparison. As *PSMD9* encodes a proteasomal subunit previously linked to PI resistance^22^, its silencing via DNA hypermethylation suggests a potential novel epigenetic mechanism of PI resistance similar to the previously reported epigenetic PSMD5 silencing^8^. After relapse to the BCMA CAR T therapy and cevostamab, the patient received talquetamab (**Figure 1A**). Taking into account that sampling was conducted prior to talquetamab therapy initiation, PBMCs exhibited 89% *GPRC5D* DNA methylation and bone marrow samples ranged from 47% to 50% (chr12: 12944100-12953200 9.1 kbp, 106 CpGs). The *GPRC5D* expression ranged from 88 TPM to 46 TPM indicating no epigenetic reason for a poor prognosis to talquetamab. Additionally, in patient 2, hypermethylation was observed in the *CRBN* gene, (**Figure 2B**), an epigenetic aberration we recently linked to IMiD resistance^19^. Before undergoing BLMF treatment, this patient had undergone six cycles of Rd in the third line and Pd in the seventh line (**SI Figure 1**). The methylation degree of the intragenic *CRBN* region overlapping with two enhancers was 60% pre-BLMF, which remained stable at 62% post-BLMF (chr3:3172800-3174401, 1.6 kbp, 20 CpGs). The patient also acquired a *CUL4B* c.2170G>A (p.E724K) mutation (VAF: 0.25, **SI Table 3**), within the ROC1 binding site, located in proximity to other *CUL4B* mutations previously reported in IMiD-resistant MM patients^10^. To assess whether *BCMA* hypermethylation was haplotype-specific and to evaluate the potential contribution of hydroxymethylation, we performed phased analysis of 5mC and 5hmC profiles across key loci including *BCMA, FCRL5, CRBN*, and *PSMD9* (**SI Figure 5**). Methylation patterns were consistent between haplotypes with no evidence of allele-specific differences, indicating that both alleles acquired hypermethylation. Furthermore, 5hmC levels were uniformly low across all examined loci, indicating that hydroxymethylation does not contribute to regulation at these regions.

**Figure 2.**
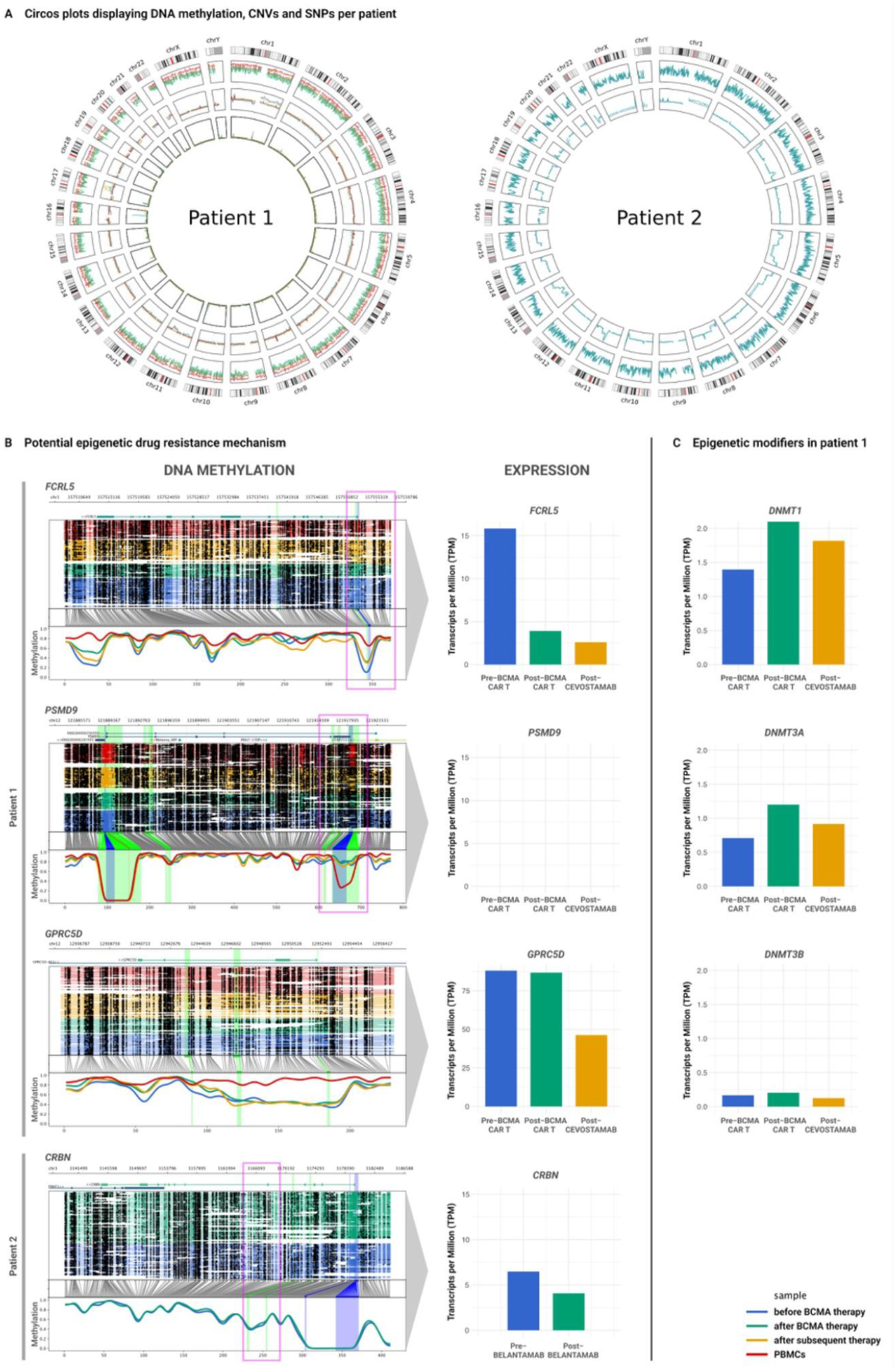
(Facing page): Circos plots illustrating multi-layered genomic data and epigenetic alterations alongside gene expression levels linked to resistance mechanisms to prior or subsequent treatments. (**A**) Circos plots display, from outer to inner layers, (epi)genome-wide DNA methylation, coverage/CNAs, and SNVs. Different samples are color-coded. **(B)** DNA methylation changes in regulatory regions are indicated by pink brackets. Regulatory regions are colour-coded: promoters in blue and enhancers in green. In patient 1, an increase in *FCRL5* promoter methylation was observed in the post-CAR T sample, which persisted in the post-cevostamab sample and was associated with downregulation of FCRL5 expression. This patient was primarily refractory to cevostamab but showed a favorable response to talquetamab. *GPRC5D* remained unmethylated at all time points and though decreased, it was still well expressed in the post-cevostamab/pre-talquetamab sample. Additionally, patient 1 exhibited hypermethylation in a 3’ regulatory region overlapping the *PSMD9* gene, which may be implicated in PI resistance and warrants further investigation. In patient 2, hypermethylation of an intragenic *CRBN* region overlapping two enhancers was observed, a modification previously linked to IMiD resistance. **(C)** Modest upregulation of the DNA methyltransferases DNMT1, DNMT3A, and DNMT3B in the post-CAR T sample compared to the pre-CAR T sample in patient 1.

## Discussion

Next-generation sequencing (NGS) technologies like whole-genome sequencing (WGS) and whole-exome sequencing (WES) have significantly enhanced our understanding of MM immunotherapy resistance^7,9,14,15^. Despite growing evidence that epigenetic dysregulation plays a crucial role in resistance to standard therapies such as IMiDs and PIs^8,19^, its role in immunotherapy resistance remains largely unexplored. Prior studies have only examined epigenetic mechanisms in the context of *GPRC5D-*directed therapies^13,17^, leaving BCMA- and FCRL5-directed immunotherapies uninvestigated. In this study, we systematically investigated all potential layers of tumor-intrinsic treatment escape, particularly focusing on the DNA methylome, by leveraging the latest technical advancements in Oxford Nanopore Technologies. Unlike conventional short-read NGS approaches such as Illumina or Ion Torrent, ONT enables direct, simultaneous detection of whole-(epi)genome-wide SVs, SNVs, DNA methylation and hydroxymethylation in a single sample, without the need for sample splitting to conduct several workflows or preprocessing steps like bisulfite conversion. This long-read sequencing technology excels in resolving complex genomic landscapes, including large SVs and repetitive regions, making it especially valuable in MM research where bone marrow aspirates often contain limited tumor cell numbers. Whole-genome coverage and CNA profiling via ONT can also serve as critical quality control measurement for assessing sample purity (**Figure 3, SI Figure 3**). By providing a comprehensive, multi-layered view of treatment resistance, long-read ONT sequencing overcomes traditional sequencing limitations, represents an integrated streamlined approach, and facilitates a deeper understanding of clonal evolution.

**Figure 3.**
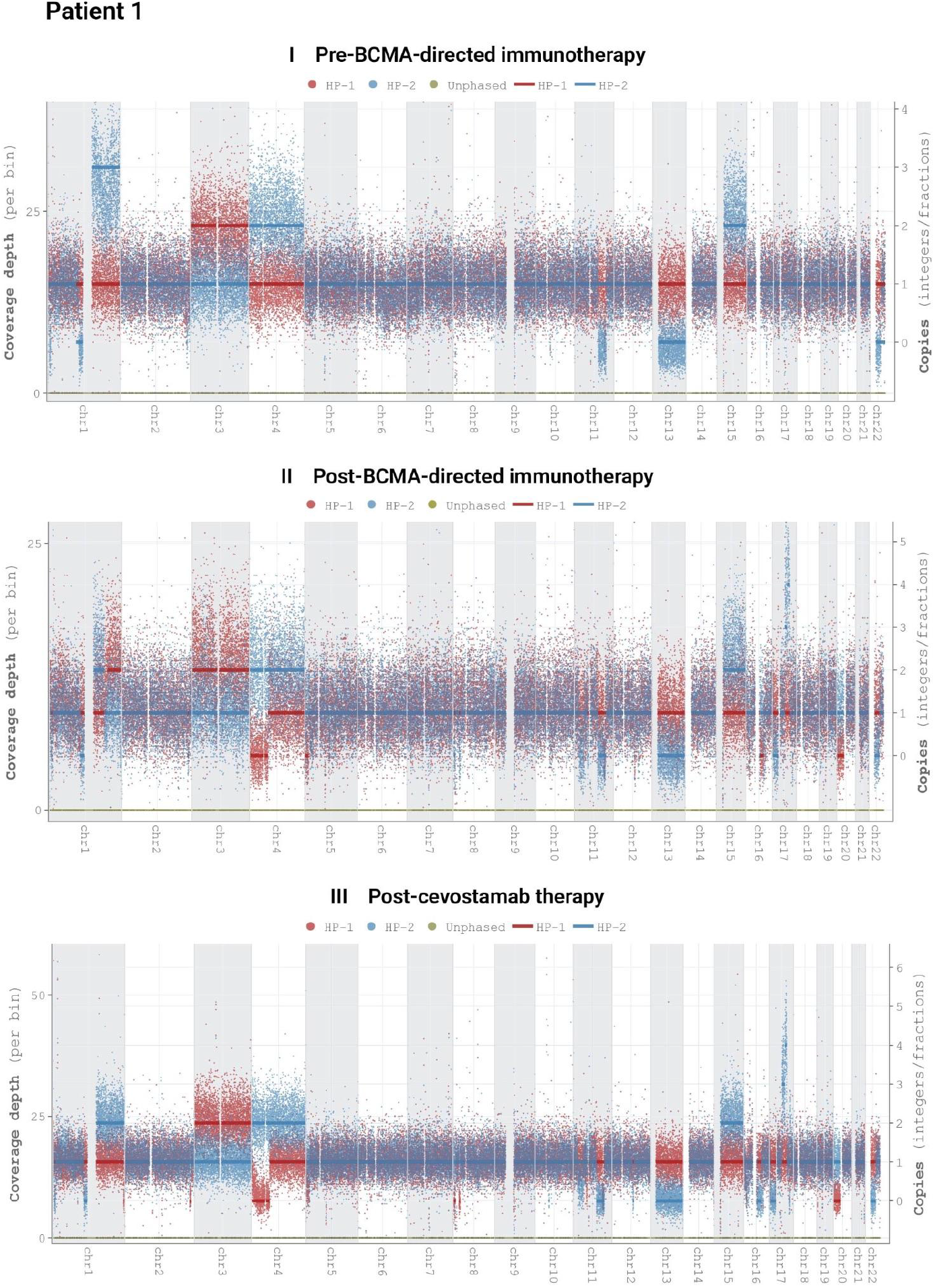
(Facing page): CNA analysis by whole genome coverage for Patient 1. Coverage depth for each phased haplotype pair is shown on the left of each graph. Combined coverage demonstrates whole-genome sequencing depth across all samples exceed 20x. Patient 1 exhibits extensive chromosomal alterations across all three timepoints including chromosome gains (+1q, +3, +15) and chromosomal losses (del 11q, del 13), consistent with a complex karyotype typical of MM. Meanwhile, the *BCMA* locus at 16p remained unaffected, indicating that *BCMA* methylation calling (**Figure 1B** and **SI Figure 7**) is independent of CNAs and coverage depth. Corresponding CNA analysis data for Patient 2 are available in **SI Figure 3**. Multiple myeloma-related target genes affected by CNAs in both patients are detailed in **SI Table 4**.

In our cases, MM was generally characterized by global hypomethylation, consistent with previous reports^18^. However, intriguingly, drug resistance in MM appears to be associated with a gain of methylation at distinct genomic loci, including acquired hypermethylation of regulatory regions of drug targets. In one patient who relapsed after BCMA CAR T-cell therapy, we detected extensive hypermethylation spanning the entire *BCMA* gene, accompanied by transcriptional downregulation and a complete loss of *BCMA* expression on the cell surface. These findings identify epigenetic silencing of *BCMA* as a novel resistance mechanism to BCMA-directed CAR T-cell therapy, as no CNAs or SNVs were detected in or overlapping with the *BCMA* gene. Further analysis confirmed that this hypermethylation was an acquired phenomenon rather than a pre-existing trait, as it was absent in MM patients naive to BCMA-targeting treatments. Notably, read-level methylation analysis revealed that pre-treatment reads segregated into two distinct patterns -one methylated and one unmethylated-indicating the coexistence of two clonal populations. Post-treatment, all reads displayed a uniformly methylated pattern, consistent with selective outgrowth of a pre-existing methylated subclone under CAR T-cell pressure (**SI Figure 7**). On the other hand, its presence in PBMCs emphasizes its crucial role as a regulator of gene expression. Interestingly, at the time of relapse, the same patient exhibited hypermethylation of the *FCRL5* promoter, accompanied by significant downregulation of its expression. Given the absence of genetic alterations in *FCRL5*, this epigenetic modification may be causal for the patient’s primary resistance to subsequent, unsuccessful FCRL5-directed cevostamab therapy. Taken together, these findings highlight the role of DNA methylation and epigenetic plasticity as key drivers of resistance to multiple immunotherapies in MM. A particularly noteworthy observation was the upregulation of DNA methyltransferases (DNMTs) in the relapse sample, specifically *DNMT1, DNMT3A*, and *DNMT3B* (**Figure 2C**). DNMT1 is primarily responsible for maintaining existing methylation patterns at hemimethylated CpG sites during DNA replication, while DNMT3A may be involved in RNA methylation. DNMT3B plays a well-known role in establishing *de novo* DNA methylation marks^23^. The increased expression of these enzymes may suggest a potential association with the acquired hypermethylation of *BCMA* and *FCRL5*, as well as the broader global hypermethylation observed in the post-treatment sample. However, additional studies would be necessary to confirm a causal relationship and not all immunotherapy target genes were affected by aberrant methylation, for instance, *GPRC5D* remained unmethylated, and the patient responded well to subsequent talquetamab treatment. Phased 5mC and 5hmC data indicates that BCMA and other examined loci do not exhibit meaningful haplotype-specific differences or locus-specific 5hmC enrichment, suggesting that hydroxymethylation is unlikely to contribute to regulation at these regions in this context. However, we identified a persistent epigenetic resistance memory of earlier treatments, e.g. we recently identified *PSMD5* promoter hypermethylation as a novel epigenetic mechanism of resistance to PIs^8^. In patient 1, hypermethylation of another *PSMD* gene, *PSMD9*, was observed, which may similarly contribute to PI resistance. PSMD9 has been reported to function as a chaperone involved in the assembly of the 19S subunit of the proteasome, assisting in proteasomal substrate degradation^24^. Lower expression levels of 19S subunit genes (*inter alia PSMD9*) have been correlated with increased PIs resistance in a meta-analysis of BTZ-resistant MM patients, as well as *in vitro* studies of various cancer cell lines. Although the underlying mechanism is not fully understood, it has been suggested that reduced expression of 19S subunit genes may increase the level of active 20S proteasomes, enhancing tolerance to PIs by triggering a cytoprotective stress response that mitigates PI-induced proteotoxic stress^22^. In patient 2 a hypermethylation of an intragenic *CRBN* region was detected, a modification we recently associated with IMiD resistance^19^. The persistence of epigenetic resistance memory has critical implications for MM treatment strategies. If tumor cells retain an epigenetic imprint from previous therapies, this could shape their responsiveness to subsequent treatments, potentially leading to cross-resistance between different immunotherapies. Future studies should investigate whether epigenetic modifications induced by one therapy influence the efficacy of others, particularly in the context of sequential BCMA-, FCRL5-, and GPRC5D-directed therapies. We anticipate that comprehensive genetic, epigenetic, and transcriptomic profiling of the target will be crucial for selecting the most promising therapy for each patient individually and for early detection of emerging resistant subclones. Additionally, larger patient cohorts will be required to determine whether SVs, SNVs, or DNA hypermethylation are the predominant mechanisms by which tumor cells lose *BCMA, FCRL5* and *GPRC5D* expression. Understanding the interplay between genetic and epigenetic resistance mechanisms may pave the way for novel therapeutic strategies, including the use of hypomethylating agents or DNMT inhibitors to restore antigen expression and resensitize tumor cells to immunotherapy. As MM treatment paradigms continue to evolve, integrating epigenetic profiling into clinical decision-making could provide a more comprehensive approach to overcoming therapy resistance and improving patient outcomes.

### Summary

In conclusion, our study provides the first evidence that acquired DNA hypermethylation serves as a resistance mechanism to BCMA- and FCRL5-directed immunotherapies. By elucidating the interplay between epigenetic modifications and antigen escape, we highlight the need to integrate epigenetic profiling into resistance/relapse studies. The examples of persistent epigenetic resistance mechanisms derived from earlier treatments (such as *PSMD9* and *CRBN* enhancer methylation) suggest that methylation marks act as memory traces of a cancer cell’s developmental and treatment history. Changing therapy alters the clonal selection pressure, potentially but not necessarily eliminating resistant clones from previous treatments, reinforcing the importance of repetitive testing. These findings pave the way for personalized treatment strategies and future research into targeted epigenetic therapies to overcome resistance in multiple myeloma patients.

## Data Availability

All data produced in the present study are available upon reasonable request to the authors

https://doi.org/10.5281/zenodo.15168660

## Acknowledgments

The authors acknowledge the Genomics Unit of the Fundacion Parque Cientifico de Madrid for its technical expertise and assistance with RNA-sequencing and express their gratitude to the patients who participated in this study.

## Disclosure of Conflicts of Interest

J.M-L. is an equity shareholder of Altum Sequencing Co. F.B. received honoraria and/or travel/accommodation expenses from BMS, AbbVie and Janssen. T.D. is an equity shareholder of Oxford Nanopore Technologies. The remaining authors declare no competing financial interests.

## Data Availability Statement

The Nanopore and RNA Seq data is available at the European Genome-phenome Archive (EGA) (https://ega-archive.org) under the project accession number XXXXXX. Source code for the analysis pipeline is available at https://doi.org/10.5281/zenodo.15168660. All requests for raw and analyzed data and materials will be promptly replied. Any data and materials that can be shared will be released via a Material Transfer Agreement. For requests, contact L.H. via e-mail at or T.D. at.

## Sources of Funding

L. Haertle was funded by the Deutsche Forschungsgemeinschaft (DFG, German Research Foundation) project ID: 493951700 and 544189139 and the CRIS cancer foundation. S. Barrio was funded by the Instituto de Salud Carlos III (FIS No. PI21/00314 and Miguel Servet CP22/00082). J.M.-L. was funded by Instituto de Salud Carlos III (ISCIII) and was co-funded by the European Union and the CRIS Cancer Foundation. F.B. was funded by the DFG TRR 387/1 (project ID: 514894665) and DFG BA 2851/7-1 (project ID: 537477296). The CNIO Bioinformatics Unit is supported by project IMPaCT-Data (IMP/00019) funded by the Agencia Estatal de Investigacion, Instituto de Salud Carlos III (ISCIII), co-funded by FEDER, “A way of making Europe’’; the project PID2021-124188NB-I00 funded by MCIN/AEI /10.13039/501100011033 and by the European Union, ERDF “A way of making Europe”, by Comunidad de Madrid (P2022/BMD-7379, iTIRONET) co-financed by European Structural and Investment Fund, the EOSC4Cancer project funded by the Horizon Europe Framework Programme under grant agreement number 101058427; the PRYCO234528VALI project funded by the Fundación científica de la Asociación Española Contra el Cáncer, the project code grant HR23-00051 funded by “la Caixa” foundation; project PMP22/00064, Instituto de Salud Carlos III (ISCIII), and funded by the European Union, the Recovery, Transformation and Resilience Plan (PRTR) through Next Generation EU recovery funds (MRR).

## Notes

### Funding Statement

L. Härtle was funded by the Deutsche Forschungsgemeinschaft (DFG, German Research Foundation) project ID: 493951700 and 544189139 and the CRIS cancer foundation. S. Barrio was funded by the Instituto de Salud Carlos III (FIS No. PI21/00314 and Miguel Servet CP22/00082). J.M.-L. was funded by Instituto de Salud Carlos III (ISCIII) and was co-funded by the European Union and the CRIS Cancer Foundation. F.B. was funded by the DFG TRR 387/1 (project ID: 514894665) and DFG BA 2851/7-1 (project ID: 537477296). The CNIO Bioinformatics Unit is supported by project IMPaCT-Data (IMP/00019) funded by the Agencia Estatal de Investigacion, Instituto de Salud Carlos III (ISCIII), co-funded by FEDER, "A way of making Europe"; the project PID2021-124188NB-I00 funded by MCIN/AEI /10.13039/501100011033 and by the European Union, ERDF "A way of making Europe", by Comunidad de Madrid (P2022/BMD-7379, iTIRONET) co-financed by European Structural and Investment Fund, the EOSC4Cancer project funded by the Horizon Europe Framework Programme under grant agreement number 101058427; the PRYCO234528VALI project funded by the Fundación científica de la Asociación Española Contra el Cáncer, the project code grant HR23-00051 funded by "la Caixa" foundation; project PMP22/00064, Instituto de Salud Carlos III (ISCIII), and funded by the European Union, the Recovery, Transformation and Resilience Plan (PRTR) through Next Generation EU recovery funds (MRR).

### Author Declarations

Clinical Research Ethics Committee (20/326) of the Biomedical Research Institute of the Hospital 12 de Octubre gave ethical approval for this work.

### Summary of Updates

Data from one patient whose post-treatment sample had low tumor infiltration (3%) were removed. The manuscript now presents two patients with high-quality sequential samples and confirmed tumor-specific copy number alterations by whole-genome Nanopore sequencing. A read-level methylation heterogeneity analysis of the BCMA promoter (36 CpG sites) was added. Pre-CAR T reads segregated into two distinct methylation patterns, while post-CAR T and post-cevostamab reads showed a single uniformly hypermethylated pattern, demonstrating selective outgrowth of a pre-existing subclone rather than gradual methylation gain. Haplotype-phased analysis of 5-methylcytosine and 5-hydroxymethylcytosine at key loci (BCMA, FCRL5, CRBN, PSMD9) was expanded, confirming bi-allelic epigenetic silencing and uniformly low 5hmC levels (<5%), ruling out active TET-mediated demethylation. RNA-seq data showing detectable CD38 transcripts (2.7 TPM) in the pre-CAR T sample were added, confirming that the absence of surface CD38 protein was consistent with epitope masking from prior daratumumab rather than transcriptional loss. Genomic analyses were expanded with comprehensive SNV and CNA/SV data from both Nanopore and targeted tchDNA-Seq, confirming no genetic alterations in BCMA or FCRL5 and identifying acquired mutations at relapse including BRAF p.V640E and CUL4B p.E724K. An error referencing DNMT2 instead of DNMT3A in the Discussion was corrected. Figure 3 was replaced with whole-genome CNA landscape plots.

## References

1. Lin, Y. et al. Idecabtagene vicleucel for relapsed and refractory multiple myeloma: post hoc 18-month follow-up of a phase 1 trial. Nat. Med. 29, 2286–2294 (2023).

2. Berdeja, J. G. et al. Ciltacabtagene autoleucel, a B-cell maturation antigen-directed chimeric antigen receptor T-cell therapy in patients with relapsed or refractory multiple myeloma (CARTITUDE-1): a phase 1b/2 open-label study. Lancet Lond. Engl. 398, 314–324 (2021).

3. Legato, L. et al. Mechanisms of Resistance to CAR T-Cells and How to Overcome Them. Methods Protoc. 8, 108 (2025).

4. Dimopoulos, M. A. et al. Belantamab Mafodotin, Pomalidomide, and Dexamethasone in Multiple Myeloma. N. Engl. J. Med. 391, 408–421 (2024).

5. Lesokhin, A. M. et al. Elranatamab in relapsed or refractory multiple myeloma: phase 2 MagnetisMM-3 trial results. Nat. Med. 29, 2259–2267 (2023).

6. Rasche, L., Hudecek, M. & Einsele, H. CAR T-cell therapy in multiple myeloma: mission accomplished? Blood 143, 305–310 (2024).

7. Lee, H. et al. Mechanisms of antigen escape from BCMA- or GPRC5D-targeted immunotherapies in multiple myeloma. Nat. Med. 29, 2295–2306 (2023).

8. Haertle, L. et al. Single-Nucleotide Variants and Epimutations Induce Proteasome Inhibitor Resistance in Multiple Myeloma. Clin. Cancer Res. Off. J. Am. Assoc. Cancer Res. 29, 279–288 (2023).

9. Truger, M. S. et al. Single- and double-hit events in genes encoding immune targets before and after T cell-engaging antibody therapy in MM. Blood Adv. 5, 3794–3798 (2021).

10. Barrio, S. et al. IKZF1/3 and CRL4CRBN E3 ubiquitin ligase mutations and resistance to immunomodulatory drugs in multiple myeloma. Haematologica 105, e237–e241 (2020).

11. Haertle, L. et al. Clonal competition assays identify fitness signatures in cancer progression and resistance in multiple myeloma. HemaSphere 8, e110 (2024).

12. Mi, X., Penson, A., Abdel-Wahab, O. & Mailankody, S. Genetic Basis of Relapse after GPRC5D-Targeted CAR T Cells. N. Engl. J. Med. 389, 1435–1437 (2023).

13. Derrien, J. et al. Acquired resistance to a GPRC5D-directed T-cell engager in multiple myeloma is mediated by genetic or epigenetic target inactivation. Nat. Cancer 4, 1536–1543 (2023).

14. Da Vià, M. C. et al. Homozygous BCMA gene deletion in response to anti-BCMA CAR T cells in a patient with multiple myeloma. Nat. Med. 27, 616–619 (2021).

15. Samur, M. K. et al. Biallelic loss of BCMA as a resistance mechanism to CAR T cell therapy in a patient with multiple myeloma. Nat. Commun. 12, 868 (2021).

16. Munawar, U. et al. Impaired FADD/BID signaling mediates cross-resistance to immunotherapy in Multiple Myeloma. Commun. Biol. 6, 1299 (2023).

17. Ma, S. et al. Genetic and epigenetic mechanisms of GPRC5D loss after anti-GPRC5D CAR T-cell therapy in multiple myeloma. Blood J. blood.2024026622 (2025) doi:10.1182/blood.2024026622.

18. Duran-Ferrer, M. et al. The proliferative history shapes the DNA methylome of B-cell tumors and predicts clinical outcome. Nat. Cancer 1, 1066–1081 (2020).

19. Haertle, L. et al. Cereblon enhancer methylation and IMiD resistance in multiple myeloma. Blood 138, 1721–1726 (2021).

20. Patiño-Escobar, B., Ramos, R., Linares, M., Mejía, A. & Alcalá, S. CD38: From Positive to Negative Expression after Daratumumab Treatment. Cureus 12, e7627 (2020).

21. Perez de Acha, O. et al. CD38 antibody re-treatment in daratumumab-refractory multiple myeloma after time on other therapies. Blood Adv. 7, 6430–6440 (2023).

22. Tsvetkov, P. et al. Suppression of 19S proteasome subunits marks emergence of an altered cell state in diverse cancers. Proc. Natl. Acad. Sci. U. S. A. 114, 382–387 (2017).

23. Law, J. A. & Jacobsen, S. E. Establishing, maintaining and modifying DNA methylation patterns in plants and animals. Nat. Rev. Genet. 11, 204–220 (2010).

24. Kaneko, T. et al. Assembly pathway of the Mammalian proteasome base subcomplex is mediated by multiple specific chaperones. Cell 137, 914–925 (2009).

