## Supplementary Material for "Multi-Omic Profiling Reveals Epigenetic Drivers of Immunotherapy Resistance in Multiple Myeloma"

### Supplementary Information

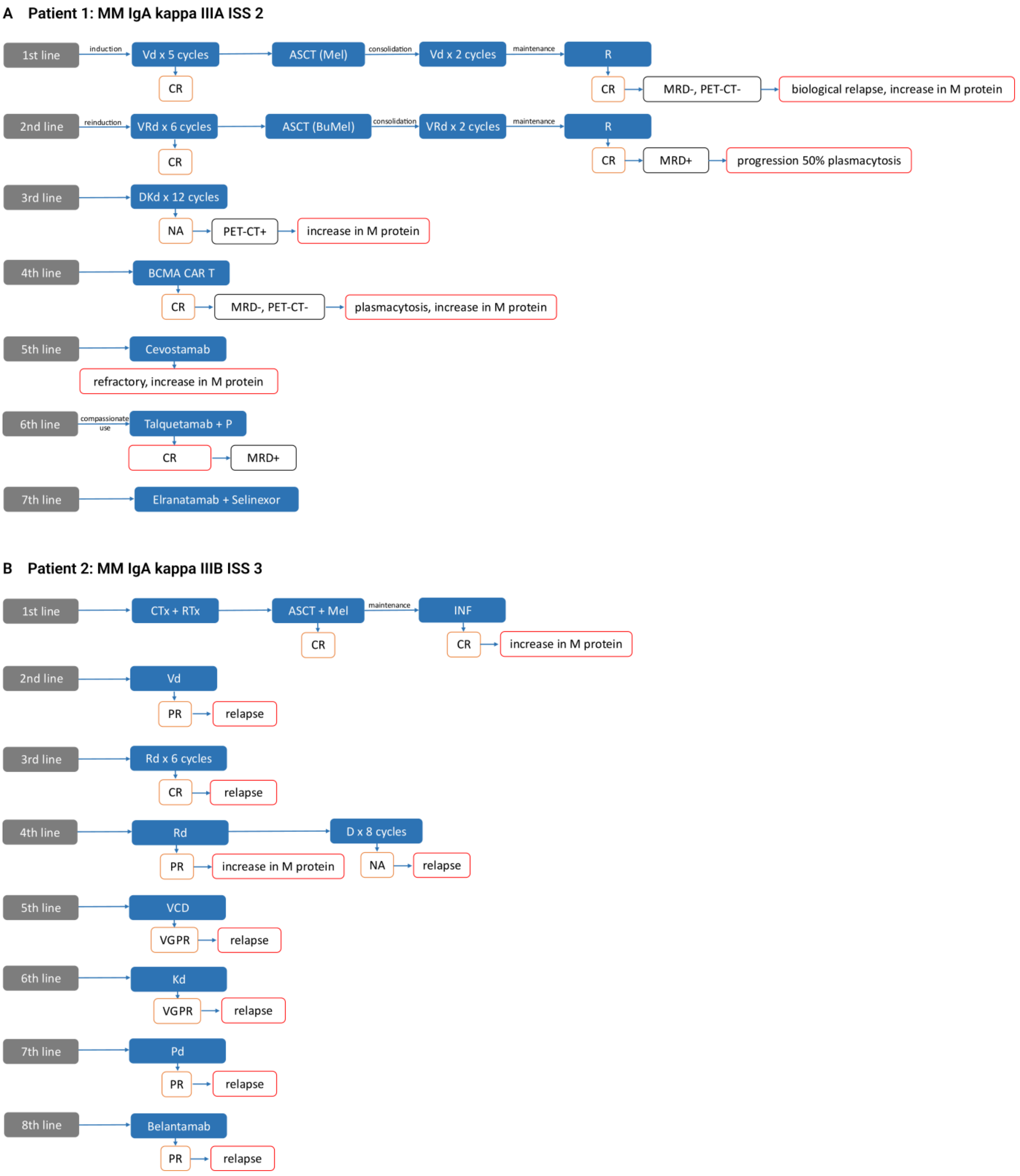


**Supplementary figure 1: Detailed treatment scheme and patients' responses under study.**
(A) patient 1 and (B) patient 2.

Abbreviations: ASCT, autologous hematopoietic stem cell transplantation; BJ, Bence-Jones; Bu, busulfan; CR, complete response; CTx, chemotherapy; d, dexamethasone; D, daratumumab; FC, flow cytometry; Ig, immunoglobulin; INF, interferon; ISS, international staging system; K, carfilzomib; KT, kidney transplant; M protein, monoclonal protein; Mel, melphalan; MRD, minimal residual disease; NA, not available; P, pomalidomide; PET-CT, positron emission tomography-computed tomography; PR, partial response; R; lenalidomide; RT, radiotherapy; RTx, radiotherapy; T, thalidomide; V, bortezomib; VGPR, very good partial response; VenVd, bortezomib-venetoclax-dexamethasone.

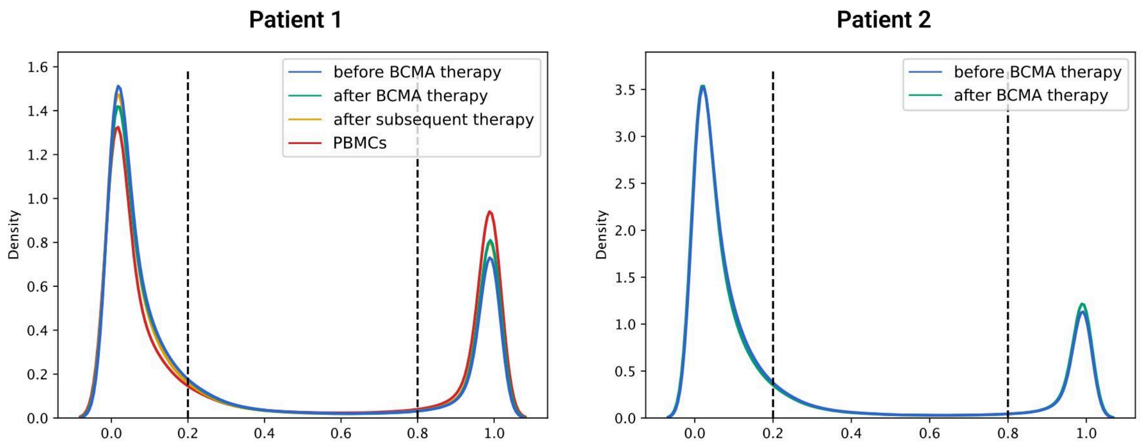


**Supplementary figure 2: Global DNA hypomethylation in MM with focal hypermethylation linked to treatment resistance.**

Global DNA methylation analysis displays that all MM samples were hypomethylated compared to their PBMC controls, however, there was an increase in hypermethylated loci in the post-BCMA immunotherapy sample, compared to the pre-sample, particularly in patient one. Methylation degrees lower than 20% were considered “unmethylated”, methylation over 80% “methylated”, respectively. Plots were generated with the scoredist option of Methylartist.

.

### Patient 2

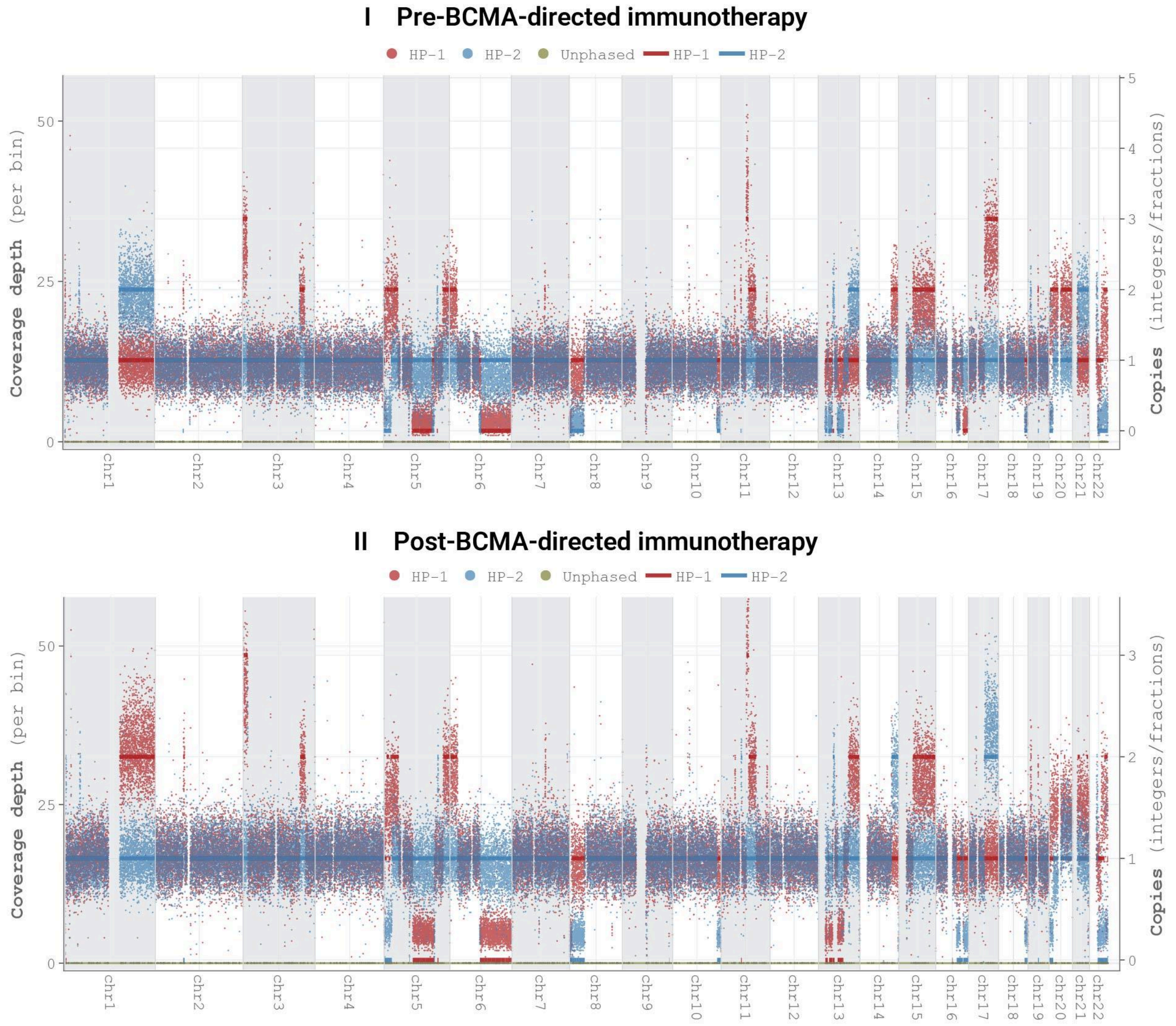


**Supplementary figure 3: CNA analysis by whole genome coverage for Patient 2**
Coverage depth for each phased haplotype pair is shown on the left of each graph. Combined coverage demonstrates whole-genome sequencing depth across all samples exceed 20x. Patient 2 exhibits a complex genomic architecture with extensive segmental CNAs spanning large chromosomal regions, generally below whole arm or chromosome level, except for the canonical +1q amplification. The CNA profile remains mostly stable across treatment timepoints. Multiple myeloma-related target genes affected by CNAs are detailed in **SI Table 4.**

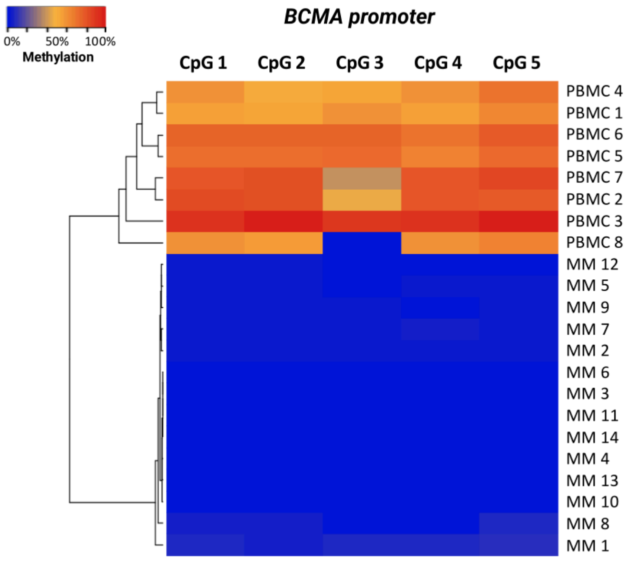


**Supplementary figure 4: *BCMA* promoter methylation in MM patients naïve to BCMA immunotherapy and PBMCs.**

The DBS heatmap presents individual samples as rows and single CpG sites as columns. An unmethylated state is depicted in blue, while a methylated state appears in red. *BCMA* promoter methylation was absent in MM patients who had not received BCMA-targeting immunotherapy. In contrast, PBMCs from all patients exhibited methylation. This indicates that *BCMA* expression is regulated via DNA methylation.

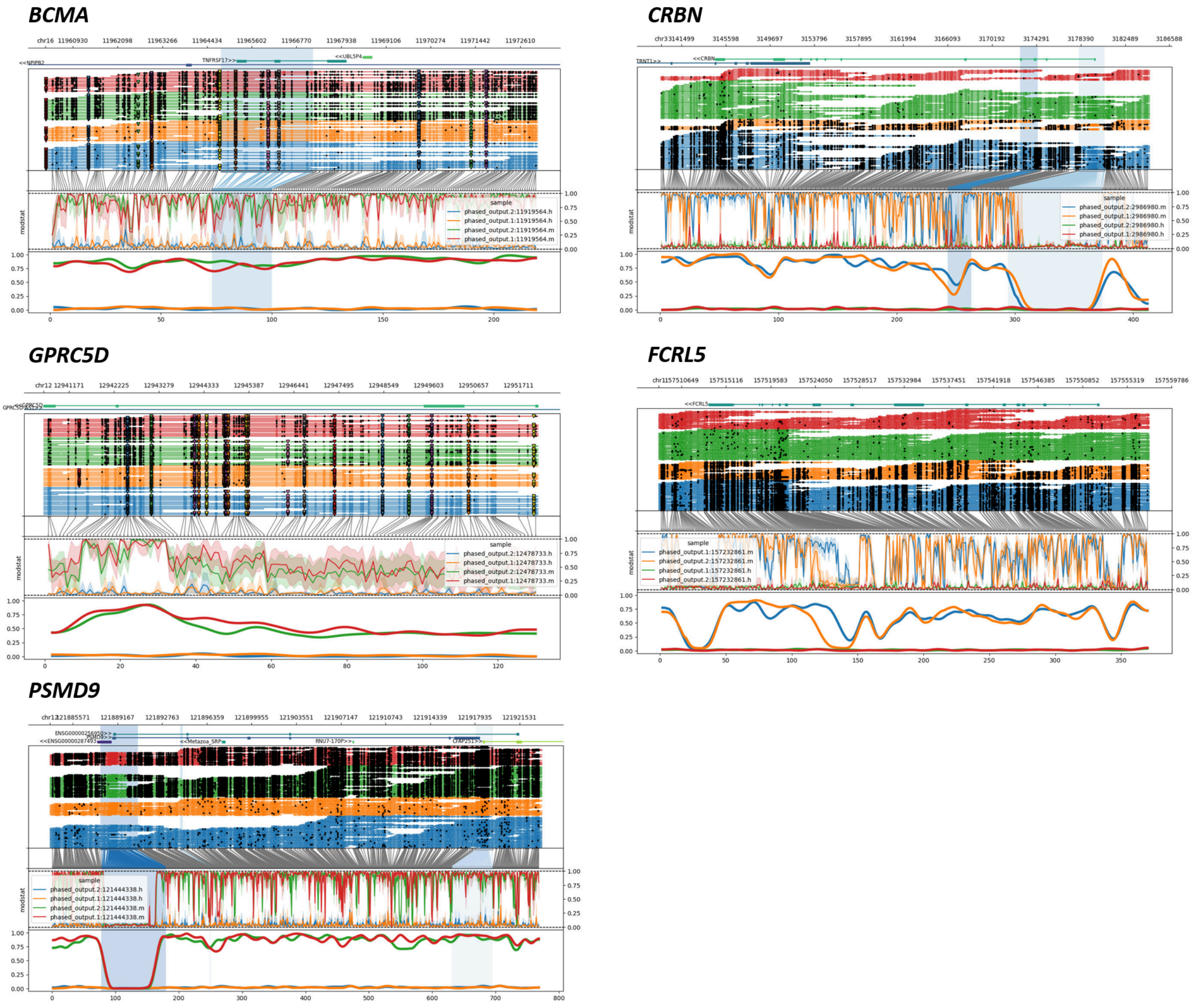


**Supplementary figure 5: Phased 5mC and 5hmC profiles at representative loci in an exemplary sample.**

Shown are phased 5-methylcytosine (5mC) and 5-hydroxymethylcytosine (5hmC) profiles for different loci of interest in one representative sample. Reads are grouped and color-coded by haplotype. Germline variants are indicated by black triangles within the reads; no somatic variants were detected. Methylation levels (modstat) are displayed separately for 5mC and 5hmC for each haplotype. Overall, this analysis shows no significant 5hmC enrichment or haplotype-specific differences in methylation at the investigated loci.

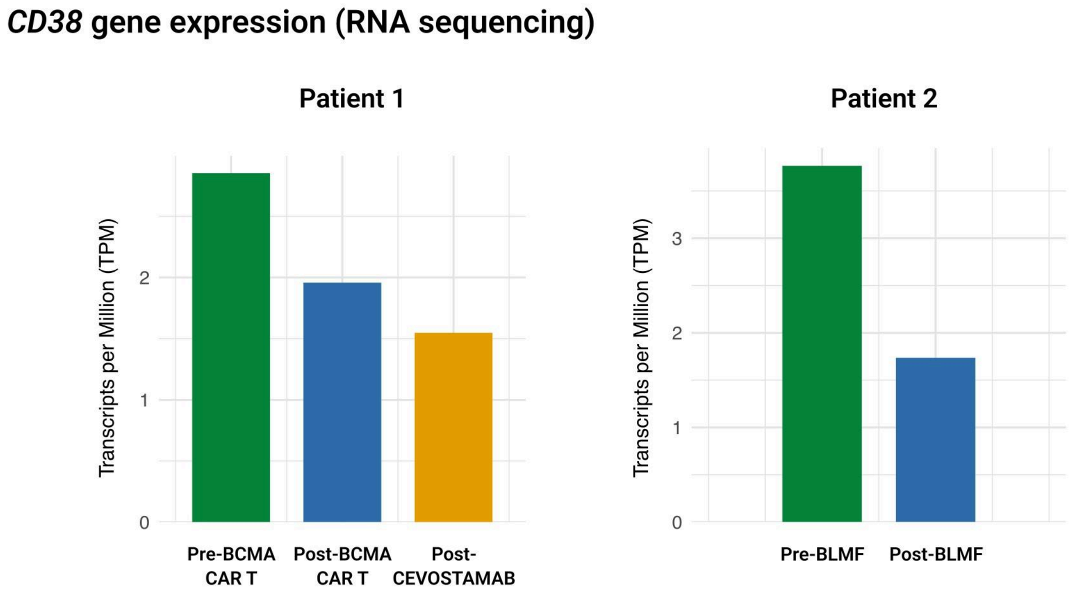


**Supplementary figure 6: *CD38* transcript expression across treatment timepoints.**
RNA-seq quantification of *CD38* expression (Transcripts per Million, TPM) in sequential bone marrow samples. In patient one, *CD38* transcripts were detectable at all timepoints (2.85 TPM pre-BCMA CAR T, 1.96 TPM post-BCMA CAR T, 1.55 TPM post-cevostamab), despite the absence of CD38 surface protein detection by flow cytometry in the pre-CAR T sample (Figure 1D). This confirms that the lack of surface CD38 was not due to transcriptional silencing. In patient two, *CD38* expression decreased from 3.76 TPM pre-BLMF to 1.73 TPM post-BLMF.

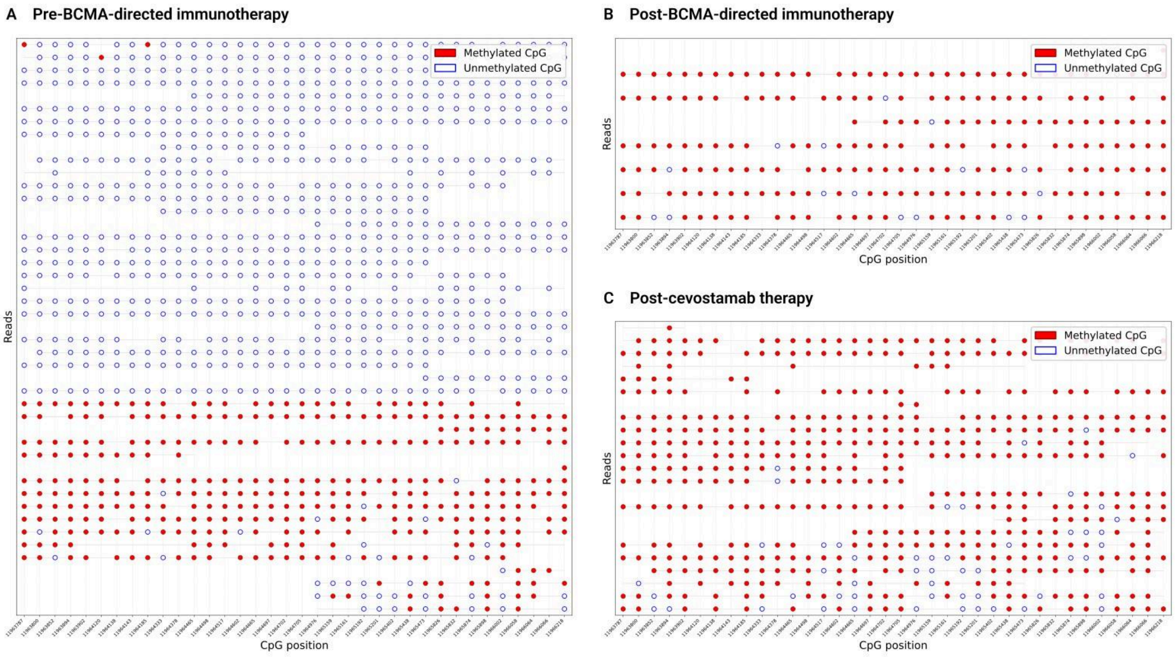

**Supplementary figure 7: Read-level *BCMA* promoter methylation patterns reveal clonal selection under therapeutic pressure.**

Methylation heterogeneity analysis of the *BCMA* promoter region (36 CpG dinucleotides) in patient one across sequential timepoints. Each row represents an individual sequencing read; filled circles indicate methylated CpGs, open circles indicate unmethylated CpGs. (A) Pre-CAR T: 45 reads segregate into 2 distinct methylation patterns, indicating the presence of two clonal populations, one methylated and one unmethylated. (B) Post-CAR T: All 7 reads display a single, uniformly methylated pattern, consistent with clonal selection of the methylated population under CAR T-cell pressure. (C) Post-cevostamab: All 23 reads maintain the same methylated pattern, indicating persistence of the resistant clone.

**Supplementary table 1:** Clinical characteristics of patients with multi-refractory MM.

| Parameter | P1 | P2 |
|---|---|---|
| **Age at diagnosis, years** | 40-45 | 60-65 |
| **MM subtype according to the secreted Ig** | IgA κ | IgA κ |
| **Durie Salmon** | IIIA | IIIB |
| **ISS Stage** | ISS 2 | ISS 3 |
| **Total lines of therapies** | 6 | 8 |
| **Cytogenetics** | tri(13) | not available |
| **CAR T therapy** | yes | no |
| **Triple class refractory** (IMiDs, PIs, anti-CD38 antibody) | yes | yes |
| **Quadruple-drug refractory,** resistant to at least one IMiDs (Lenalidomide, Thalidomide and/or Pomalidomide), two PIs (Bortezomib and Carfilzomib), and an anti-CD38 antibody (Daratumumab) | yes | yes |

Abbreviations: Ig, immunoglobulin; IMiD, immunomodulatory drugs; ISS, international staging system; P, patient; PI, proteasome inhibitor.

**Supplementary table 2:** Primers used for DBS and pyrosequencing.

| Gene | Primer | Template-specific sequence (5'-3') | Amplicon length | Chromosomal assay location[c] | Number of CpGs | Annealing Temp. [°C] |
|---|---|---|---|---|---|---|
| **DBS** | | | | | | |
| *BCMA* Promoter | F<br>R | [a]TGTATTTTGTAATTGTTATTTGGTTGAGAA<br>[b]TAAAAACACTACCCAACCATCTAC | 354 bp | Chr16:+ 11965000:11965353:1 | 5 | 57 °C |
| **Pyro** | | | | | | |
| *BCMA* Assay 1 | F<br>R<br>S | TGTATTTTGTAATTGTTATTTGGTTGAGAA<br>[Biotin]TAAAAACACTACCCAACCATCTAC<br>ATTAGATGTGGTATTTAAATTTTTA | 354 bp | Chr16:+ 11965000-11965353 | 3 | 56°C |
| *BCMA* Assay 2 | F<br>R<br>S | AGTTTGAGGTTAGTTTGGTTAATTT<br>[Biotin]CCTTTCACTAAATTAATCACACCTTTATA<br>GGAGGTTGTAGTGAG | 314 bp | Chr16:+ 11965903-11966216 | 3 | 52°C |

[a] upstream universal adapter 1: ACACTCTTTCCCTACACGACGCTCTTCCGATCT

[b] upstream universal adapter 2: GTGACTGGAGTTCAGACGTGTGCTCTTCCGATCT

[c] Ensembl release GRCh38p12

Abbreviations: Chr, chromosome; F, forward primer; R, reverse primer; S, sequencing primer; +, sense strand.

**Supplementary table 3:** Results of the targeted capture-hybridization DNA sequencing panel (tchDNA-Seq).

| Pat. | Time point | Gene | Chr. | Position | Ref. Allele | Var. Allele | Depth | Ratio | Transcript | cDNA position | Prot. position |
|---|---|---|---|---|---|---|---|---|---|---|---|
| **1** | Pre-CAR T | - | - | - | - | - | - | - | - | - | - |
| | Post-CAR T | *FAT3* | chr11 | 92352333 | A | T | 578 | 0.27 | NM_001367949.2 | c.221A>T | p.D74V |
| | Post-cevo | *PRDM1* | chr6 | 106099450 | A | G | 340 | 0.05 | NM_001198.4 | c.562A>G | p.K188E |
| | | *FAT3* | chr11 | 92352333 | A | T | 489 | 0.52 | NM_001367949.2 | c.221A>T* | p.D74V |
| | | *DIS3* | chr13 | 72761699 | G | A | 315 | 0.05 | NM_014953.5 | c.2458C>T | p.R820W |
| **2** | Pre-bela | *DIS3* | chr13 | 72777446 | G | C | 191 | 0.44 | NM_014953.5 | c.628C>G* | p.R210G |
| | Post-bela | *BRAF* | chr7 | 140753336 | A | T | 369 | 0.15 | NM_001374258.1 | c.1919T>A | p.V640E |
| | | *DIS3* | chr13 | 72777446 | G | C | 207 | 0.49 | NM_014953.5 | c.628C>G* | p.R210G |
| | | *CUL4B* | chrX | 120534577 | C | T | 132 | 0.25 | NM_001079872.2 | c.2170G>A | p.E724K |

All SNVs were classified as missense variants. *possible SNP

Abbreviations: bela, belantamab mafodotin; cevo, cevostamab; Chr, Chromosome; Pat, patient; Prot, protein; Ref, reference; Var, variant.

**Supplementary table 4:** Multiple Myeloma-related target genes affected by copy number alterations.

| Gene | Chr | Start | End | Copy number state | | | | |
|---|---|---|---|---|---|---|---|---|
| | | | | Patient 1 | | | Patient 2 | |
| | | | | Pre-CAR T | Pos-CAR T | Post-cevo | Pre-bela | Post-bela |
| *PSMA5* | chr1 | 109399041 | 109426448 | 1 | 1 | 1 | 2 | 2 |
| *PSMD4* | chr1 | 151254708 | 151267479 | 4 | 3 | 3 | 3 | 3 |
| *PSMB4* | chr1 | 151399559 | 151401937 | 4 | 3 | 3 | 3 | 3 |
| *FCRL5* | chr1 | 157513376 | 157552515 | 4 | 3 | 3 | 3 | 3 |
| *SLAMF7* | chr1 | 160739056 | 160754821 | 4 | 3 | 3 | 3 | 3 |
| *PSMD1* | chr2 | 231056844 | 231173116 | 2 | 1 | 1 | 2 | 2 |
| *CRBN* | chr3 | 3144627 | 3179727 | 3 | 3 | 3 | 4 | 4 |
| *PSMD6* | chr3 | 64010549 | 64024010 | 3 | 3 | 3 | 2 | 2 |
| *PSMD2* | chr3 | 184299197 | 184309050 | 3 | 3 | 3 | 2 | 2 |
| *CD38* | chr4 | 15778274 | 15853232 | 3 | 2 | 2 | 2 | 2 |
| *ABCG2* | chr4 | 88090149 | 88231628 | 3 | 3 | 3 | 2 | 2 |
| *PSMG4* | chr6 | 3231402 | 3303373 | 2 | 2 | 2 | 3 | 3 |
| *PSMB1* | chr6 | 170535119 | 170553307 | 2 | 2 | 2 | 1 | 1 |
| *PSMC3* | chr11 | 47418768 | 47426473 | 2 | 1 | 2 | 2 | 2 |
| *PSMC1* | chr14 | 90256526 | 90275429 | 2 | 2 | 2 | 3 | 3 |
| *PSMA4* | chr15 | 78540404 | 78552417 | 3 | 3 | 3 | 3 | 3 |
| *IGF1R* | chr15 | 98648538 | 98964530 | 3 | 3 | 3 | 3 | 3 |
| *PSMB6* | chr17 | 4796143 | 4798502 | 2 | 1 | 1 | 2 | 2 |
| *PSMC5* | chr17 | 63827151 | 63832026 | 2 | 2 | 2 | 4 | 3 |
| *PSMD12* | chr17 | 67337915 | 67366605 | 2 | 2 | 2 | 3 | 3 |
| *PSMF1* | chr20 | 1113239 | 1189415 | 2 | 1 | 1 | 1 | 1 |
| *DNMT3B* | chr20 | 32762384 | 32809359 | 2 | 2 | 2 | 3 | 2 |
| *PSMA7* | chr20 | 62136732 | 62143440 | 2 | 2 | 2 | 3 | 2 |
| *PSMG1* | chr21 | 39174768 | 39183488 | 2 | 2 | 2 | 3 | 3 |
| *XBP1* | chr22 | 28794554 | 28800597 | 1 | 1 | 1 | 1 | 1 |

Abbreviations: bela, belantamab mafodotin; cevo, cevostamab; Chr, Chromosome.

**Supplementary table 5:** *BCMA* pyrosequencing results of cell line triplicates (MM1S, NCI-H929, Amo1, OPML, OPM2, KMS11).

| Sample | *BCMA* Pyro Assay 1 | | | | | *BCMA* Pyro Assay 2 | | | | |
|---|---|---|---|---|---|---|---|---|---|---|
| | CpG 1 | CpG 2 | CpG 3 | Mean Meth. [%] | Mean Meth. Tripl. [%] | CpG 1 | CpG 2 | CpG 3 | Mean Meth. [%] | Mean Meth. Tripl. [%] |
| MM1S R 1 | 2.49 | 0.00 | 4.20 | **2.23** | | 2.77 | 2.97 | 1.92 | **2.55** | |
| MM1S R 2 | 2.51 | 0.00 | 3.22 | **1.91** | | 3.34 | 1.73 | 0.00 | **1.69** | |
| MM1S R 3 | 2.33 | 0.00 | 3.43 | **1.92** | **2.02** | 2.75 | 0.00 | 1.68 | **1.48** | **1.91** |
| NCI-H929 R 1 | 2.96 | 0.00 | 4.80 | **2.59** | | 0.00 | 0.00 | 0.00 | **0.00** | |
| NCI-H929 R 2 | 0.00 | 0.00 | 5.32 | **1.77** | | NA | NA | NA | **NA** | |

| | | | | | | | | | | |
|---|---|---|---|---|---|---|---|---|---|---|
| NCI-H929 R 3 | 2.34 | 0.00 | 2.99 | **1.78** | **2.05** | NA | NA | NA | **NA** | **0.00** |
| Amo1 R 1 | 2.42 | 1.40 | 4.92 | **2.91** | | 2.94 | 0.00 | 0.00 | **0.98** | |
| Amo1 R 2 | 2.05 | 0.00 | 4.08 | **2.04** | | 4.53 | 0.00 | 0.00 | **1.51** | |
| Amo 1 R 3 | 2.43 | 0.00 | 3.83 | **2.09** | **2.35** | 5.66 | 0.00 | 1.96 | **2.54** | **1.68** |
| OPM2 R 1 | 2.34 | 1.71 | 3.41 | **2.49** | | 5.87 | 0.00 | 0.00 | **1.96** | |
| OPM2 R 2 | 3.27 | 1.77 | 3.65 | **2.90** | | 2.63 | 0.00 | 1.79 | **1.47** | |
| OPM2 R 3 | 2.57 | 1.64 | 3.78 | **2.66** | **2.68** | 3.74 | 0.00 | 0.00 | **1.25** | **1.56** |
| KMS11 R 1 | 2.77 | 1.79 | 3.69 | **2.75** | | 4.95 | 2.50 | 0.00 | **2.48** | |
| KMS11 R 2 | NA | NA | NA | **NA** | | 3.33 | 0.00 | 0.00 | **1.11** | |
| KMS11 R 3 | 1.88 | 1.45 | 2.25 | **1.86** | **2.31** | 3.21 | 2.58 | 2.41 | **2.73** | **2.11** |

Abbreviations: meth, methylation; NA, not available; pyro, pyrosequencing; R, replicate; tripl, triplicates.

### Supplementary Methods

#### Annotation sources

Genomic sequence and annotations were obtained from the GENCODE project, version 45. Regulatory regions were obtained from the Ensembl Regulatory Build, release 113. All genomic coordinates refer to the Genome Reference Consortium's human reference genome version 38 (GRCh38).

#### Bioinformatic Analysis of Nanopore Sequencing

Nanopore sequencing data was processed using a custom-developed pipeline tailored to simultaneously analyze SNVs, SVs, and DNA methylation. The pipeline was developed using the Snakemake[1] workflow management system and packages from the bioconda[2] and conda-forge[3] projects to facilitate distribution and reproducibility.

*Base Calling and Quality Control*

Raw nanopore signal data (POD5 files) was base-called using Oxford Nanopore's Dorado software[4] (v0.5.0) with the "dna_r10.4.1_e8.2_400bps_sup@v4.3.0_5mCG_5hmCG@v1" base calling model capable of detecting 5mCG and 5hmCG modifications. Base-called reads were filtered for quality score ≥9 to ensure high-quality sequence data. Quality control metrics were assessed using PycoQC[5] (v2.5.2), which provided read length distributions, quality scores, and sequencing yield statistics.

*Sequence Alignment and Processing*

Filtered reads were aligned to the GRCh38 human reference genome using Minimap2[6] (v2.26) with parameters optimized for ONT data (-x map-ont). Aligned reads were sorted and indexed using SAMtools[7] (v1.17). Only primary alignments were retained for downstream analyses by filtering out supplementary and secondary alignments (SAMtools flags "-F 2308). Coverage statistics were calculated using both SAMtools coverage and Mosdepth[8] (v0.3.8) to ensure adequate genome-wide representation.

*Methylation Analysis*

DNA methylation was analyzed using Modkit[9] (v0.5.0) for genome-wide methylation profiling. CpG methylation patterns were extracted directly from the native DNA reads without bisulfite conversion, leveraging ONT's ability to detect modified bases from electrical signal variations. Methylation levels at each CpG site were quantified as the percentage of reads showing the modification. Visualization of methylation patterns at specific genomic regions of interest was performed using Methylartist[10] (v1.3.1).

*Structural Variant (SV) Detection*

SVs were detected using Sniffles[11] (v2.3.3) with parameters tuned to identify both germline and somatic/mosaic variants (--mosaic --mosaic-include-germline), and Severus[12] (v1.2) with default parameters. This allowed detection of deletions, insertions, duplications, inversions, and translocations across the genome.

*Single Nucleotide Variant (SNV) Analysis*

For SNVs, Clair3[13] (v1.0.8) was employed with the r1041_e82_400bps_sup_v420 model specifically trained for ONT data. For samples with matched normal controls, ClairS[14] (v0.4.3) was utilized to identify somatic mutations, allowing the differentiation between germline polymorphisms and somatic variants. Phasing of variants was performed using WhatsHap[15] to determine haplotype-specific alterations.

*Copy Number Aberration (CNA) Analysis*

CNAs were analyzed using Wakhan[16] (v0.2.0) for chromosome-scale somatic profiling with default tumor-normal mode parameters, including optional parameters to exclude sex chromosomes (--contigs chr1-22) and generate copy number state lists for targeted genes (--user-input-genes).

**RNA Sequencing and bioinformatic analysis**

To quantify the concentration and determine the purity of the RNA, we used a NanoDrop ND-1000 spectrophotometer (Thermo Fisher Scientific) and a 2100 Bioanalyzer with the RNA 6000 Nano kit (Agilent). Libraries were prepared using the TruSeq stranded mRNA kit (Illumina), single-end sequencing was conducted on a NextSeq 500 (Illumina) using the High Output kit. RNA sequencing data was processed using a pipeline developed in the CNIO Bioinformatics Unit (https://github.com/cnio-bu/cluster_rnaseq) for quality control, preprocessing, alignment and quantification. The pipeline makes extensive use of Snakemake's integration with the conda package manager, to automatically take care of software requirements and dependencies.

*Quality Control and Preprocessing*

Raw data quality was assessed with FastQC[17] (v1.18) and contamination screening performed with fastq-screen[18] (v0.15.2). Adapter sequences were removed using BBDuk[19] (v39.01) with default adapter files.

*Sequence Alignment*

Filtered reads were aligned to the GRCh38 human reference genome using STAR[20] (v2.7.8a) with their respective primary assembly annotation.

*Expression Quantification*

Read alignment data were processed to generate count matrices across all samples, using featureCounts[21] (v2.0.6) with the previously described annotation. These data were used to calculate and visualize the proportion of transcripts in each sample measured as Transcripts Per Million (TPM) using a custom R script (v4.3.3).

**Flow Cytometry of BCMA expression**

The expression of *BCMA* in MM plasma cells was evaluated by incubating 50-100 µl of bone marrow patient samples with the antibody panel CD38-FITC, BCMA-PE, CD45-PerCPCy5.5, CD138-PECy7, CD56-APC and CD19-APCCy7 (Biolegend) at 4 ºC for 30 minutes in the dark. Erythrocyte lysis was then performed with ACK Lysing Buffer (Lonza). After two washing steps with PBS, samples were acquired using a FACS Canto II flow cytometer (BD Biosciences) with FACSDiva v.8.0.1 software (BD Biosciences), and data was analyzed using FlowJo v10 software (BD Biosciences).

**Deep Bisulfite Sequencing (DBS) of Patient Samples and Pyrosequencing of Cell Lines**

The primers for the DBS *BCMA* assay and pyrosequencing assay 1 were similar, with only one difference: the DBS primers were tagged with 5' universal adapters for Illumina sequencing, while the reverse pyrosequencing primer had a 5' biotin tag. Pyrosequencing assay 2 was located 550 bp downstream of assay 1, but also within the *BCMA* promoter region (**SI table 2**). DNA isolation was performed with the AllPrep DNA/RNA Micro Kit (Qiagen) and bisulfite conversion with the EpiTect Bisulfite Kit. Pyrosequencing was conducted on a PyroMark Q96 MD system (Qiagen) using the PyroMark Gold Q96 CDT Reagent Kit (Qiagen) and 10 pmol of the sequencing primer. Data analysis was performed using the Pyro Q-CpG software (Qiagen). DBS libraries were prepared as previously described[22] and paired-end sequenced on the MiSeq platform (Illumina). FASTQ files were aligned using the Amplikyzer2[23] pipeline .

**Targeted Capture-Hybridization DNA Sequencing Panel (tchDNA-Seq) for MM**

The MM-specific targeted capture-hybridization DNA sequencing panel (tchDNA-Seq) was designed, applied, validated, and bioinformatically analyzed as previously detailed[24]. This panel encompasses over 40 genes implicated in MM development and progression, drug resistance, and targets for immunotherapy. It also includes canonical loci covering IGH rearrangements, known germline polymorphisms, and frequent translocations. The panel was designed to screen for genetic alterations to aid in prognostic risk stratification and allows simultaneous assessment of copy number and single nucleotide variations. Additionally, it enables the detection of smaller insertions, deletions, translocations, and IGH rearrangements.


1. Mölder, F. *et al.* Sustainable data analysis with Snakemake. *F1000Research* **10**, 33 (2021).
2. Grüning, B. *et al.* Bioconda: sustainable and comprehensive software distribution for the life sciences. *Nat. Methods* **15**, 475–476 (2018).
3. Conda-Forge Community. The conda-forge Project: Community-based Software Distribution Built on the conda Package Format and Ecosystem. Preprint at https://doi.org/10.5281/ZENODO.4774217 (2015).
4. nanoporetech/dorado. Oxford Nanopore Technologies (2026).
5. Leger, A. & Leonardi, T. pycoQC, interactive quality control for Oxford Nanopore Sequencing. *J.*

*Open Source Softw.* **4**, 1236 (2019).

6. Li, H. New strategies to improve minimap2 alignment accuracy. *Bioinformatics* **37**, 4572–4574 (2021).
7. Danecek, P. *et al.* Twelve years of SAMtools and BCFtools. *GigaScience* **10**, giab008 (2021).
8. Pedersen, B. S. & Quinlan, A. R. Mosdepth: quick coverage calculation for genomes and exomes. *Bioinformatics* **34**, 867–868 (2018).
9. nanoporetech/modkit. Oxford Nanopore Technologies (2026).
10. Cheetham, S. W., Kindlova, M. & Ewing, A. D. Methylartist: tools for visualizing modified bases from nanopore sequence data. *Bioinformatics* **38**, 3109–3112 (2022).
11. Smolka, M. *et al.* Detection of mosaic and population-level structural variants with Sniffles2. *Nat. Biotechnol.* **42**, 1571–1580 (2024).
12. Keskus, A. G. *et al.* Severus detects somatic structural variation and complex rearrangements in cancer genomes using long-read sequencing. *Nat. Biotechnol.* 1–11 (2025) doi:10.1038/s41587-025-02618-8.
13. Zheng, Z. *et al.* Symphonizing pileup and full-alignment for deep learning-based long-read variant calling. *Nat. Comput. Sci.* **2**, 797–803 (2022).
14. Zheng, Z. *et al.* ClairS: a deep-learning method for long-read somatic small variant calling. 2023.08.17.553778 Preprint at https://doi.org/10.1101/2023.08.17.553778 (2023).
15. Martin, M. *et al.* WhatsHap: fast and accurate read-based phasing. 085050 Preprint at https://doi.org/10.1101/085050 (2016).
16. Ahmad, T. *et al.* Wakhan: reconstruction of chromosome-scale copy number profiles of tumor genomes with long-read sequencing. 2025.12.11.25342098 Preprint at https://doi.org/10.64898/2025.12.11.25342098 (2025).
17. Babraham Bioinformatics - FastQC A Quality Control tool for High Throughput Sequence Data. https://www.bioinformatics.babraham.ac.uk/projects/fastqc/.
18. Wingett, S. & Bottoms, C. StevenWingett/FastQ-Screen: FastQ Screen v0.15.3. Zenodo

https://doi.org/10.5281/zenodo.7795529 (2023).

19. Bushnell, B. BBMap: A Fast, Accurate, Splice-Aware Aligner. https://escholarship.org/uc/item/1h3515gn (2014).

20. Dobin, A. *et al.* STAR: ultrafast universal RNA-seq aligner. *Bioinformatics* **29**, 15–21 (2013).

21. Liao, Y., Smyth, G. K. & Shi, W. featureCounts: an efficient general purpose program for assigning sequence reads to genomic features. *Bioinformatics* **30**, 923–930 (2014).

22. Haertle, L. *et al.* Hypermethylation of the non-imprinted maternal MEG3 and paternal MEST alleles is highly variable among normal individuals. *PloS One* **12**, e0184030 (2017).

23. Rahmann, S. *et al. Amplikyzer: Automated Methylation Analysis of Amplicons from Bisulfite Flowgram Sequencing*. https://peerj.com/preprints/122v2 (2013) doi:10.7287/peerj.preprints.122v2.

24. Rosa-Rosa, J. M. *et al.* NGS-Based Molecular Karyotyping of Multiple Myeloma: Results from the GEM12 Clinical Trial. *Cancers* **14**, 5169 (2022).
